# CFO: Calibration-Free Odds Bayesian Designs for Dose Finding in Clinical Trials

**DOI:** 10.1101/2024.07.26.24311051

**Authors:** Jialu Fang, Wenliang Wang, Guosheng Yin

**Affiliations:** Department of Statistics and Actuarial Science, The University of Hong Kong

**Keywords:** Bayesian design, Dose-finding trial, Late-onset toxicity, Maximum tolerated dose, Phase I trial

## Abstract

The calibration-free odds type (CFO-type) of designs, as data-driven decision-making Bayesian approaches, leverage historical cumulative data across various dose levels, primarily aiming at identifying the maximum tolerated dose (MTD). Inheriting the ideas from game theory or “tug-of-war”, CFO mimics the games of force: one pushes the dose down while the other pushes it up. Extensive simulations validate that CFO-type designs maintain an optimal balance between efficiency and safety in MTD identification, with performance metrics that are comparable to, or occasionally surpass the state-of-the-art methods. This article primarily introduces the R package **CFO** for implementing and assessing CFO-type designs in phase I clinical trials. Besides, we propose integrating the mechanism of exploration and exploitation from reinforcement learning into the CFO design, leading to a novel approach: the randomized CFO (rCFO) design. The **CFO** package encompasses various variants tailored to accommodate different scenarios. Beyond the fundamental CFO design, these include the two-dimensional CFO (2dCFO) designed for drug-combination trials, accumulative CFO (aCFO) for accruing all dose information, time-to-event CFO (TITE-CFO), and fractional CFO (fCFO) which are developed to specifically address late-onset toxicity. Moreover, hybrid designs such as TITE-aCFO and f-aCFO, which integrate both late-onset toxicity and all dose information for decision making, are also included. **CFO** provides a robust set of functions used for determining subsequent cohort doses, selecting the MTD, and conducting simulations to evaluate design operating characteristics. The properties and results are presented to trial investigators through simple textual and graphical outputs. The user-friendly interface, adaptability to various design considerations, and the comprehensive implementation of CFO-type designs position **CFO** as a noteworthy tool for phase I clinical trials.

## 1 Introduction

The primary objective of a phase I clinical trial in oncology is to determine the maximum tolerated dose (MTD), which is defined as the dose at which the probability of dose-limiting toxicity (DLT) aligns with a predetermined toxicity rate.[1] Numerous dose-finding methodologies currently exist for MTD determination, categorized broadly into algorithm-based, model-based and model-assisted approaches. The 3 + 3 design,[2] a traditional algorithm-based method, has been widely employed for its simplicity and transparency. Despite these advantages, this design and its derivatives often struggle to accurately identify the MTD and tend to assign subtherapeutic doses to patients.[3] Conversely, commonly used model-based approaches, such as the continual reassessment method (CRM) [4] and the escalation with overdose control (EWOC) design,[5] accurately guide dose adjustments by continuously updating the parameters of mathematical models for dose-toxicity relationships. However, model-based designs are sensitive to the parametric model assumptions, which, if violated, may lead to poor trial performance. Model-assisted and certain algorithm-based designs, including the cumulative cohort design (CCD),[6] Bayesian optimal interval (BOIN) design,[7] the uniformly most powerful Bayesian interval (UMPBI) design,[8] and the calibration-free odds (CFO) design,[9] seek to bridge the gap between the simplicity of algorithm-based methods and the precision of model-based approaches. Unlike traditional algorithm-based designs that only use information from the current dose level, the CFO design leverages data across multiple dose levels within a Bayesian framework. This approach, being model-free or curve-free, enhances its robustness by avoiding explicit parametric assumptions about dose-toxicity relationships and intricate design parameters.

The CFO design [9] is a recently proposed novel phase I trial methodology that, as the name implies, does not require the calibration of any essential design parameters. The CFO design has demonstrated robustness, model-free, and easy to use but encounters challenges when addressing late-onset toxicity. The emergence of the TITE method and fractional method has led to the development of TITE-CFO [10] and fCFO designs for accumulating delayed toxicity. In these CFO-type designs, only a subset of the complete dose information is utilized, focusing solely on the toxicity data of the current dose position and its two neighboring positions. To utilize data at all dose levels, the accumulative CFO (aCFO) design [11] is proposed to incorporate dose information from all positions into trial decision-making. This fundamental concept of accumulation can also be applied to CFO-type designs with late-onset toxicity, such as the TITE-CFO and fCFO designs, resulting in the development of their respective extensions known as the TITE-aCFO and f-aCFO designs. [11] Recently, the 2dCFO design [12] has been developed to advance the CFO model for use in drug combination trials. Extensive simulation studies indicate comparable, and at times superior, performance compared to competing methodologies. [9, 10, 11]

All existing designs for phase I trials are greedy approaches that take a deterministic move by exploiting the past information without exploration of unknowns. To enhance the decision-making process, we propose the randomized CFO (rCFO) design, which integrates the exploration-exploitation mechanism from reinforcement learning into the CFO framework. This approach introduces a randomization scheme, similar to the multi-armed bandit problem, allowing for probabilistic dose adjustments. All the various extensions of the CFO design inherit its model-free and calibration-free nature. This characteristic significantly alleviates the burden of artificial input of design parameters, thereby enhancing the robustness and objectivity of the design.

Various related R packages have been developed for dose-finding methodologies. Examples of such packages include **BOIN** [13] and **TITEgBOIN** [14] for BOIN-type designs, **bcrm** [15] and **dfcrm** [16] for CRM-type designs, **ewoc** [17] for EWOC-type designs, and **TEQR** [18] for CCD-type designs. However, R packages for CFO-type designs are yet to be developed. In this article, we introduce a comprehensive, well-documented, and user-friendly R [19] package—the **CFO** package.

The **CFO** package, represents a comprehensive implementation of all CFO-type designs mentioned above. The package can be accessed from the Comprehensive R Archive Network, with the website address https://cran.r-project.org/package=CFO. The versatile functionalities embedded within **CFO** cover crucial aspects such as determining the subsequent cohort’s dose level, selecting the MTD for a single trial, and executing multiple simulations to obtain the operating characteristics. **CFO** offers flexibility in the choice of various CFO-type designs based on factors such as the incorporation of all dose information, the approach to handling late-onset toxicity, runtime, and whether the target drug covers a single agent or a combination dose findings. The functions utilized for distinct tasks under various CFO-type designs are illustrated in Figure 1. Beyond its functional depth, **CFO** distinguishes itself by providing a user-friendly evaluation of designs through both summary and graphical outputs via the `summary()` and `plot()` functions. This feature, while less common in other packages, is deemed valuable as it provides users with a more intuitive understanding of the model’s operational dynamics and outcomes, thereby facilitating broader utilization.

**Figure 1:**
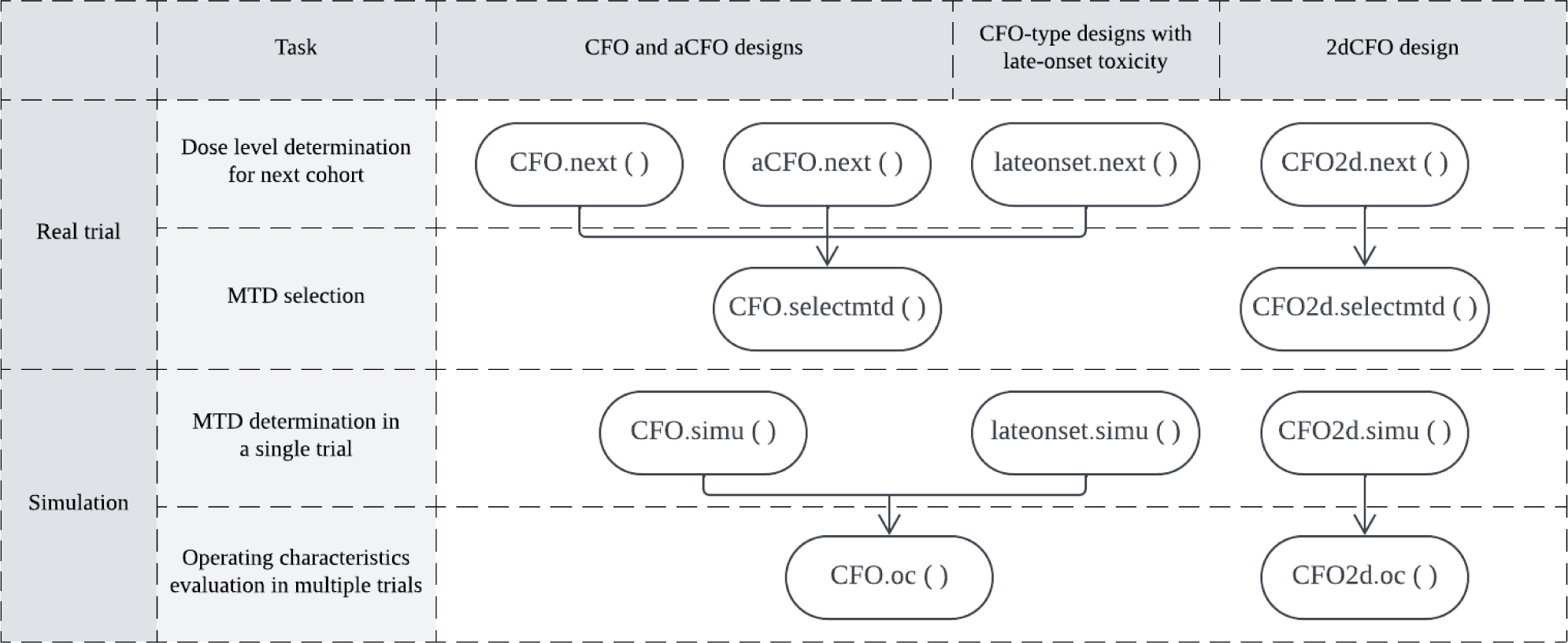
The utilization flowchart of user-visible functions in the **CFO** package.

To the best of our knowledge, this is the first R package that comprehensively implements the CFO-type designs. Section 2 will introduce all existing CFO-type designs and guide their applications in real trials. Notably, Section 2.5 offers a detailed exposition of the innovative rCFO design and presents simulation results demonstrating its effectiveness. Section 3 will instruct users on conducting single or multiple simulations in software, with results comparing them to other popular phase I designs also presented in this section. Section 4 concludes with discussions.

## 2 CFO-type designs and their application in real trials using R

In the context of toxicity monitoring, the CFO-type design aims to determine the MTD with a DLT risk probability closest to the pre-determined target rate. Figure 2 presents a summarized flowchart illustrating the sequence of steps in the CFO-type design. If the stopping conditions is not satisfied (i.e., the trial continues till exhaustion of the total sample size), for designs without late-onset toxicity, odds ratios can be calculated straightforwardly. While, for designs with late-onset toxicity, it is necessary to fill in the pending segments of data before proceeding with odds ratio calculations. With the completed or inherently complete data, the CFO-type designs make dose allocation based on the information embedded in a subset or all of the entire dose levels. This iterative process continues until the stopping condition is satisfied and the MTD is finally identified through isotonic regression. [20] The variability among CFO-type designs predominantly depends on the dose level range where the utilized information is situated, the imputing method employed for handling late-onset toxicity, and the number of target drugs involved (single or combination doses). This section will introduce different CFO-type methods and demonstrate how each CFO-type design determines the dose level for the next cohort using software. The functions available in the **CFO** package are outlined in Appendix A.1, and the meanings of the arguments within these functions are detailed in Appendix A.2.

**Figure 2:**
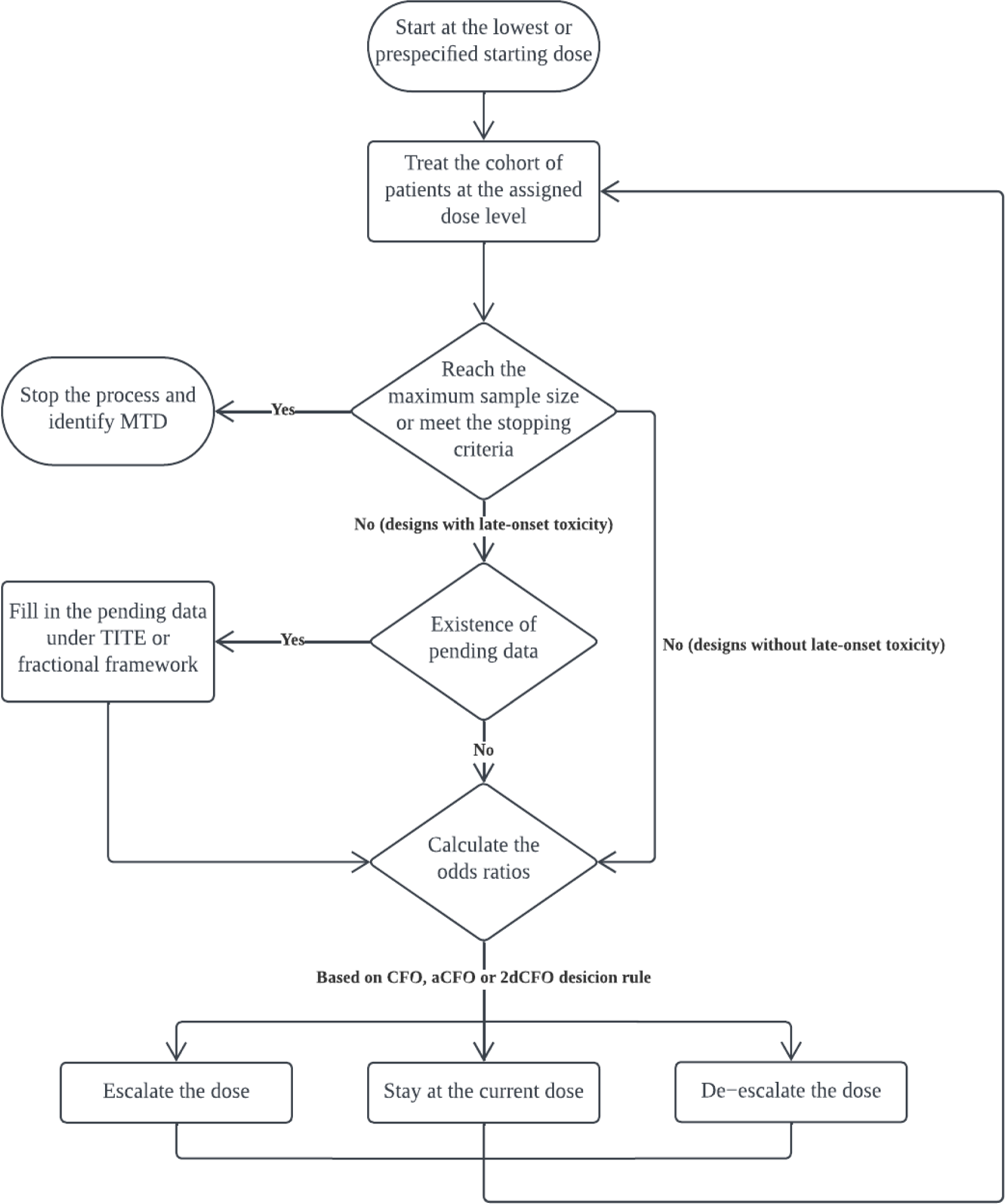
The flowchart of the Bayesian CFO-type design for phase I clinical trials.

### 2.1 The CFO design for single-drug trials

Employing a concept inspired by game theory, the CFO design determines the dose level for the next cohort of subjects by competing the current dose against its neighboring (left and right) doses, because the dose movement is typically confined to the left or right by one dose level only. [9] The clinical trial examines *K* dose levels with increasing DLT rates, *p*_1_ *< · · · < p_K_*, where *p_k_* represents the DLT probability of dose level *k*. The trial specifies a target DLT rate of *ϕ* which corresponds to the MTD. After enrolling *n* cohorts, we observe toxicity outcomes at all dose levels as 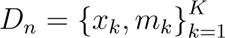, where *x_k_* and *m_k_* represent the number of observed DLTs and the number of patients treated at dose level *k*, respectively. To determine the dose level for the next cohort, the statistic denoting the odds of *p_k_ > ϕ* is defined as

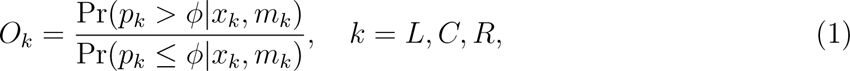

where *L, C, R* correspond to the left, current (or central) and right dose levels. The reciprocal *Ō_k_ =* 1/*O_k_* represents the odds of *p_k_ ≤ ϕ*.

In the Bayesian paradigm, the posterior distribution Pr (*p_k_ | x_k_, m_k_*) can be computed based on a beta-binomial model. Under the binomial likelihood function that *x_k_ | m_k_, p_k_ ∼* Binomial(*m_k_, p_k_*) and a beta prior distribution *p_k_ ∼* Beta(*ϕ,* 1 *− ϕ*), we apply Bayes’ rule to obtain the posterior distribution of *p_k_*,

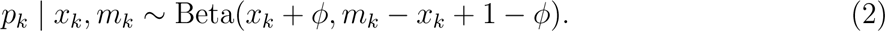

Based on the posterior distribution of *p_k_*, the odds in Equation (1) can be easily computed.

For illustrating the decision to initiate dose de-escalation, *O_C_* measures evidence of excessive toxicity at the current dose level, with a higher *O_C_* indicating a preference for dose de-escalation. Conversely, a higher *Ō_L_* reflects excessive tolerance at the left dose level, making de-escalation less favorable. The interplay between *O_C_* and *Ō_L_* results in the odds ratio *O_C_/Ō_L_*, indicating the strength of the inclination toward dose de-escalation. Similarly, *O_C_/Ō_R_* measures the inclination or strength toward escalation to the right dose level.

Adopting the Bayesian posterior distribution of the DLT rate for computing odds values incorporates consideration of the sample size. As the sample size increases, the variance of the posterior distribution diminishes, resulting in more robust evidence for making dose assignment decisions. The threshold values for the odds ratios on the left and right side of the current dose, *γ_L_* and *γ_R_*, can be pre-determined by minimizing the probability of making an incorrect decision. [9] By integrating these decision processes, the dose level for the next cohort is determined following the rule specified in Table 1. In scenarios where the current dose level is either the lowest or highest (i.e., at the boundary of the dose range), we simplify the procedure by computing the odds ratio on one side to make the decision.

**Table 1:** Decision rules for the CFO design in searching for the MTD by comparing the left- and right-side odds ratio against the threshold values *γ_L_* and *γ_R_*.

| | | $O_C/\bar{O}_L > \gamma_L$ | |
| --- | --- | --- | --- |
|  |  | Yes (De-escalation) | No (Stay) |
| $\bar{O}_C/O_R > \gamma_R$ | Yes (Escalation) | Stay | Escalation |
|  | No (Stay) | De-escalation | Stay |

In a hypothetical phase I trial with seven dose levels and a target DLT rate of 0.2, suppose the current dose level is 4. The cumulative numbers of DLTs and patients for the left, current, and right dose levels are given as (0, 1, 0) and (3, 6, 0), respectively. To decide the next cohort’s dose level, the function `CFO.next()` is executed:

~~~
R> decision <- CFO.next(target = 0.2, cys = c(0,1,0), cns = c(3,6,0),
+ currdose = 3, prior.para = list(alp.prior = 0.2, bet.prior = 0.8),
+ cutoff.eli = 0.95, early.stop = 0.95)
R> summary(decision)
~~~

The `summary()` function condenses the output into a textual summary that provides insights into the occurrence of overly toxic situations, suggests the recommended dose level for the next cohort, and determines whether the trial should be terminated.

### 2.2 The aCFO design for single-drug trials

The original CFO design makes dose allocation by examining one dose level above and one below the current dose level, while most of the existing algorithm or model-assisted designs, including interval-based designs, consider the information at the current dose level only. As the trial advances, it is crucial to consider the accumulating data at distant dose levels that could hold valuable information. This is inspired by the tug-of-war game, where even people distant from the center matter. Building upon the foundation of the CFO design, we introduce the aCFO (accumulative CFO) design, aiming at incorporating data from all available dose levels into the decision-making process. [11]

After enrolling *n* patient cohorts, all relevant dose data are combined into the cumulative dataset 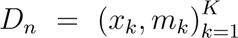. Among the *K* dose levels, the *n*th cohort is treated at dose level *C*, and there are *J* doses to the left and *H* doses to the right of *C*, satisfying *K* = *J* + *H* + 1. The DLT rates for the current dose and neighboring doses on both sides are denoted as (*p_LJ_, …, p_L_*_1_, *p_C_, p_R_*_1_, *…, p_RH_*), where *p_L_*_1_ is the DLT rate at level 1 on the left side and *p_LJ_* corresponds to level *J* on the left (i.e., *p_LJ_* = *p*_1_). Similarly, *p_R_*_1_ is the DLT rate at level 1 on the right side and *p_RH_* corresponds to level *H* on the right (i.e., *p_RH_* = *p_K_*). We use (*x_LJ_, …, x_L_*_1_, *x_C_, x_R_*_1_, *…, x_RH_*) and (*m_LJ_, …, m_L_*_1_, *m_C_, m_R_*_1_, *…, m_RH_*) to denote the respective counts of DLTs and the total numbers of patients at all dose levels. Furthermore, (*O_LJ_, …, O_L_*_1_, *O_C_, O_R_*_1_, *…, O_RH_*) and (*Ō_LJ_, …, Ō_L_*_1_, *Ō_C_, Ō_R_*_1_, *…, Ō_RH_*) can also be derived for all dose levels using Equation (1).

In the aCFO design, the concept of accumulation is employed to integrate information from all dose levels to the left (or right) of the current dose level. Building upon the odds ratios *O_C_/Ō_L_* and *O_C_/Ō_R_* defined in the previous section, two aggregated odds ratio (OR) statistics are formulated, encompassing comprehensive leftward and rightward information. These are defined by summing up individual odds ratios from the left or right side, expressed as

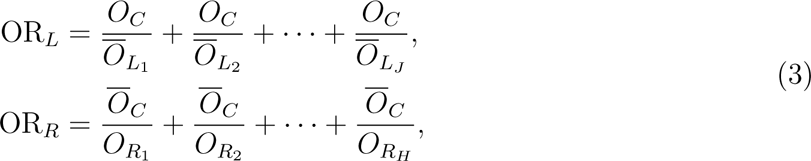

which is similar to the tug-of-war by adding up all the strengths from the left versus those from the right. The new thresholds, *γ^′^_L_* for OR*_L_* and *γ^′^_R_* for OR*_R_*, are determined by summing their respective individual thresholds, i.e., 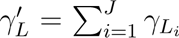 and 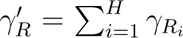. Integrating the decision processes for all these statistics yields the decision rule for the aCFO design, as outlined in Table 2.

**Table 2:**
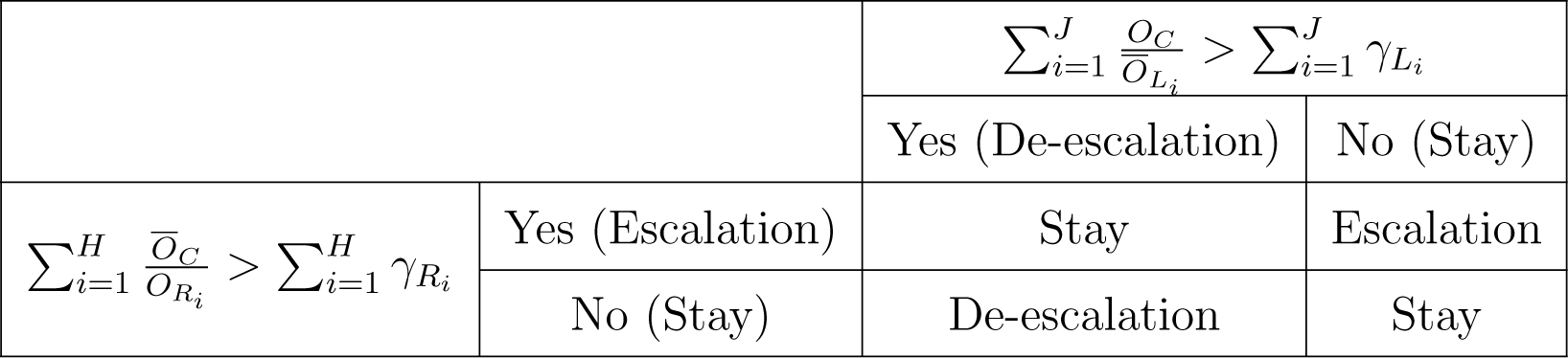
Decision rules for the aCFO design in searching for the MTD.

In contrast to the CFO design, the aCFO design leverages information from all dose levels. Extensive simulations are conducted on a 3.2 GHz Apple M1 processor with 8 cores. The result indicates that the aCFO design exhibits slightly superior performance, attributed to its ability to integrate information from a broader range of dose levels. In terms of runtime, for each trial simulation, aCFO requires approximately 0.4 seconds, whereas CFO takes around 0.15 seconds. Despite slightly longer processing time for aCFO, the inherent swiftness of CFO-type designs, owing to their calibration-free and model-free nature, allows us to ignore the impact of runtime in this context. At the beginning of a trial when no data are collected at most of the dose levels, aCFO reduces to CFO due to the simple summation in Equation (3). Within the **CFO** package, the choice between the CFO and aCFO designs depends on specific needs, considering the range of dose information to be incorporated.

The process of determining the dose level for the next cohort in the aCFO design resembles that of the CFO design. The primary distinction between `aCFO.next()` and `CFO.next()` lies in the arguments related to the existing toxicity outcomes. In `aCFO.next()`, these arguments are denoted as ays and ans, representing the accumulative numbers of DLTs and patients across all dose levels. Conversely, `CFO.next()` exclusively utilizes dose information from the left, current, and right dose levels (cys and cns). Utilizing ays and ans and following the parameter settings as in `CFO.next()`, it employs the function:

~~~
R> ays <- c(0, 0, 1, 0, 0, 0, 0); ans <- c(3, 3, 6, 0, 0, 0, 0)
R> decision <- aCFO.next(target = 0.2, ays = ays, ans = ans, currdose = 3,
+ prior.para = list(alp.prior = 0.2, bet.prior = 0.8),
+ cutoff.eli = 0.95, early.stop = 0.95)
R> summary(decision)
~~~

Both `CFO.next()` and `aCFO.next()` form the foundation for various CFO-type designs. These functions are intrinsic to the CFO package, serving as essential components for other functions within the package. For example, they are integrated into `CFO.simu()` to facilitate the simulation of a single CFO or aCFO trial. Furthermore, these functions are embedded within `lateonset.next()` and `lateonset.simu()` to execute other CFO-type designs with late-onset toxicity.

### 2.3 Designs with late-onset toxicities

The CFO and aCFO designs necessitate that toxicity outcomes for all preceding cohorts are observed before the next cohort arrives, as these outcomes inform the determination of the subsequent cohort’s dose level. They rely on binary DLT data, where a patient experiencing a DLT event is denoted as *y* = 1, and if no DLT occurs, *y* is set to 0. However, late-onset toxicity commonly arises in phase I dose-finding trials, especially for targeted therapies or immunotherapies. The follow-up time for pending data contains rich information that should be utilized to help pin down the right dose. Frameworks such as TITE and fractional schemes have been developed to address late-onset toxicity by representing pending DLT data with decimal values between 0 and 1 (i.e., partial observation of the outcome). Designs accommodating late-onset toxicity, including TITE-CFO,[10] fCFO, TITE-aCFO, and f-aCFO,[11] have been proposed under these two frameworks. Specifically, the time-to-event framework assumes that the time to DLT follows a uniform distribution, while the fractional framework employs the Kaplan–Meier estimator without making assumptions about the time-to-event data.

The assessment window is denoted by *τ*, the follow-up time of a pending-outcome patient is *u* and the time to DLT is represented by *T*. In the framework of the time-to-event weighting model, assuming a uniform distribution over the interval [0, *τ*], the TITE-CFO and TITE-aCFO designs address pending *y* by considering the expected outcome conditioned on the actual follow-up time. For a subject treated at dose level *k*, the imputed outcome is

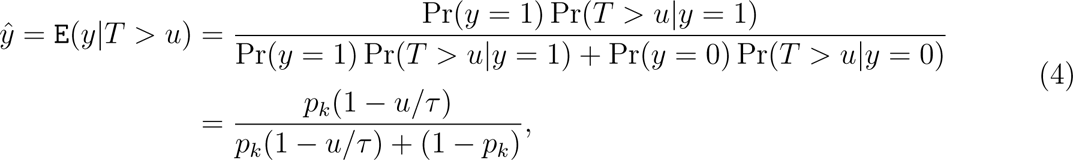

where *p_k_*represents the true DLT rate at dose level *k*.

In the fractional framework, the contribution of pending data is modeled using the Kaplan– Meier estimator. Both fCFO and f-aCFO designs handle pending *y* by estimating the conditional probability of toxicity occurrence in the remaining follow-up period, given that the toxicity event has not occurred by time *u*. This can be formulated nonparametrically as

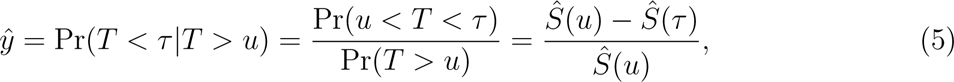

where *Ŝ*(*·*) denotes the Kaplan–Meier estimator for the survival function *S*(*·*).

For designs with late-onset toxicity, as illustrated in Figure 2, it is essential to complete the pending segments of toxicity data for all enrolled subjects prior to computing odds ratios in each iteration. Following this, the dose movement is dictated with both complete and imputed outcomes by the CFO (dose movement presented in Table 1) or aCFO (dose movement presented in Table 2) decision rule.

When addressing late-onset toxicity, users can utilize `lateonset.next()` to assign the most appropriate dose level to each enrolled cohort. Adhering to the previous setting that each cohort comprises three subjects, we assume the situation where the 7th cohort enters the trial at a recorded time of `current.t = 9.41`. The arguments `enter.times`, `dlt.times`, and `doses`—reflecting individual conditions—are consequently structured as lists with a length size of 18 (corresponding to 18 patients in the first six cohorts). The maximal assessment window size is set as three months, and other parameters persist with the pre-assumed values from Section 2.1. Taking the f-aCFO design as an example, the code below demonstrates the utilization of the function `lateonset.next()` for dose assignment to the newly enrolled cohort:

~~~
R> p.true <- c(0.01, 0.07, 0.20, 0.35, 0.50, 0.65, 0.80)
R> prior.para = list(alp.prior = 0.2, bet.prior = 0.8)
R> enter.times <- c(0, 0.266, 0.638, 1.54, 2.48, 3.14, 3.32, 4.01, 4.39, 5.38, 5.76,
+ 6.54, 6.66, 6.93, 7.32, 7.66, 8.14, 8.74)
R> dlt.times <- c(0, 0, 0, 0, 0, 0, 0, 0, 0, 0.610, 0, 2.98, 0, 0, 1.95, 0, 0, 1.48)
R> doses <- c(1, 1, 1, 2, 2, 2, 3, 3, 3, 4, 4, 4, 3, 3, 3, 4, 4, 4)
R> decision <- lateonset.next(design = ‘f-aCFO’, target = 0.2, p.true = p.true,
+ currdose = 4, assess.window = 3, enter.times = enter.times,
+ dlt.times = dlt.times, current.t = 9.41, doses = doses,
+ prior.para = prior.para, cutoff.eli = 0.95, early.stop = 0.95)
R> summary(decision)
~~~

### 2.4 The 2dCFO design for drug-combination trials

Combined drugs have become commonplace for cancer treatment. To enhance the CFO design and its robustness and precision for drug-combination trials, the 2dCFO approach was recently introduced. [12] Decision-making within the two-dimensional toxicity probability space is conducted by performing two independent one-dimensional CFO analyses along both the horizontal and vertical axes. Suppose that we study the combined toxicities of two drugs, drug A and drug B, with *J* and *K* doses respectively. Let *p_jk_* denote the joint DLT rate of dose combination (*j, k*), *j* = 1, *…, J*, *k* = 1, *…, K*. After enrolling *n* cohorts, we observe the DLT outcomes as 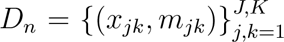, where *x_jk_* and *m_jk_* are respectively the observed number of DLTs and the total number of patients treated at dose combination (*j, k*). Let *C* denote the current dose. The four adjacent doses—*L*, *R*, *U*, and *D*, corresponding to the left, right, up, and down positions—are considered for decision-making.

The 2dCFO design uses the same odds to measure the tendency for escalating/de-escalating the dose level as its one-dimensional CFO counterpart, which is defined as

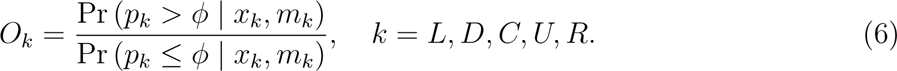

Subject to the constraints of partial ordering, decisions are based on two separate one-dimensional dose sequences. One is in the horizontal direction, denoted by *{L, C, R}*, and the other is in the vertical direction, denoted by *{D, C, U}*. Both sequences have monotonically ascending DLT rates. Additionally, the dose sequences *{L, C, U}* and *{D, C, R}* (both also have monotonically ascending DLT rates) are considered when necessary in the second stage. The comprehensive decision rules for 2dCFO are presented in Table 3.

**Table 3:** Decision rules for the 2dCFO design for finding the MTD.

| Vertical Decisions | Horizontal Decisions |  |  |
| --- | --- | --- | --- |
|  | Escalate | Stay | De-escalate |
| Escalate | Escalate to $R$ if $O_R < O_U$, escalate to $U$ if $O_R > O_U$, and randomly select $R$ or $U$ if $O_R = O_U$ | Escalate to $U$ | Next dose is decided by 1dCFO analysis on $\{L, C, U\}$ |
| Stay | Escalate to $R$ | Stay | De-escalate to $L$ |
| De-escalate | Next dose is decided by 1dCFO analysis on $\{D, C, R\}$ | De-escalate to $D$ | De-escalate to $L$ if $O_L > O_D$, de-escalate to $D$ if $O_L < O_D$, and randomly select $L$ or $D$ if $O_L = O_D$ |

The `CFO2d.next()` function can be used to determine the subsequent dose levels in a trial under the 2dCFO design. It shares an identical set of arguments in `CFO.next()` while extending certain parameters, including cys, cns, and currdose, to a two-dimensional context. Assuming that we are exploring the dose combinations of drug A and drug B, the two-element vector currdose indicates the current dose combination level for the drug-combination trial. The first element represents the dose level of drug A, and the second element represents the dose level of drug B. Let cys and cns denote the DLTs and patient allocations for both the current dose and the eight adjacent doses surrounding it within a 2D toxicity probability space. This setup encompasses the information for a total of nine doses, organized in a 3 *×* 3 matrix. Given the dose level of drug A (corresponding to a specific row), the matrix represents information for the three dose levels of drug B from low to high, reading from left to right. Similarly, given the dose level of drug B (corresponding to a specific column), the matrix represents information for the three dose levels of drug A from low to high, reading from top to bottom. The code below illustrates the practical application of the `CFO2d.next()` function:

~~~
R> cns <- matrix(c(3, 3, 0, 0, 6, 0, 0, 0, 0), nrow = 3, byrow = TRUE)
R> cys <- matrix(c(0, 1, 0, 0, 2, 0, 0, 0, 0), nrow = 3, byrow = TRUE)
R> decision <- CFO2d.next(target = 0.3, cys = cys, cns = cns,
+ currdose = c(2, 3), cutoff.eli = 0.95, early.stop = 0.95, seed = 1)
R> summary(decision)
~~~

The `CFO2d.next()` function distinguishes itself from `CFO.next()` by offering two options for escalation (drug A or drug B) and two options for de-escalation (drug A or drug B). During the initial trial phase, especially when administering low-toxicity doses where DLTs are often not observed, the 2dCFO design considers the odds of escalation for both drug A and drug B to be equivalent. Consequently, the design randomly selects one of the two drugs for escalation, assigning equal probability to each. It is recommended to set a random seed to ensure reproducibility of the results, as demonstrated in the example.

In real trials, `CFO.next()`, `aCFO.next()`, `lateonset.next()` and `CFO2d.next()` can be utilized to provide recommendations for the dose level of the next cohort. After completing the trial, which has assigned the doses to all cohorts, the next task is to select the most appropriate dose as the MTD, elaborated in Section 2.6.

### 2.5 The rCFO design with randomization for single-drug trials

Existing phase I trial designs predominantly employ greedy approaches that make deterministic decisions by exploiting the data from treated patients, without exploration of unknowns. Traditional exploiting strategies tend to focus solely on utilizing past information to maximize immediate outcomes, often leading to local optima. In contrast, an exploitation-exploration approach balances the use of existing information (exploiting) with the investigation of new possibilities (exploring), potentially leading to more optimal long-term outcomes. To better balance the trade-off between risk and reward, we propose the innovative rCFO design, which has not been previously explored in the dose-finding literature. The core idea is to incorporate the exploitation-exploration mechanism from reinforcement learning into the CFO framework. The original CFO design determines dose movement by constructing two odds ratios, *π_L_* = *O_C_/Ō_L_* and *π_R_* = *O_C_/Ō_R_*, and comparing them against thresholds *γ_L_* and *γ_R_*, respectively. In the CFO framework, dose adjustments are made deterministically, following specific rules as detailed in Table 1. The rCFO design introduces a randomization scheme, akin to the idea in multi-armed bandit problems, allowing for probabilistic dose adjustments. As delineated in Table 4, this stochastic decision rule facilitates dose escalation, de-escalation, or staying at the same dose based on calculated probabilities.

**Table 4:** Decision rules for the rCFO design in searching for the MTD.

| | | $\pi_L = O_C/\bar{O}_L > \gamma_L$ | |
| --- | --- | --- | --- |
|  |  | Yes (De-escalation) | No (Stay) |
| $\pi_R = \bar{O}_C/O_R > \gamma_R$ | Yes (Escalation) | Stay if $\pi_L = \pi_R$; otherwise, escalate with probability $\pi_L/(\pi_L + \pi_R)$ and de-escalate with probability $\pi_R/(\pi_L + \pi_R)$ | escalate with probability $\pi_R/(\pi_L + \pi_R)$ and stay with probability $\pi_L/(\pi_L + \pi_R)$ |
| | No (Stay) | de-escalate with probability $\pi_L/(\pi_L + \pi_R)$ and stay with probability $\pi_R/(\pi_L + \pi_R)$ | Stay |

The rCFO design normalizes odds ratios, *π_L_*, and *π_R_* into probabilities, thereby constructing randomization probabilities for dose escalation, de-escalation, and staying at the same dose. We conducted 5000 simulations to compare the CFO and rCFO designs under five fixed scenarios in Cheung and Chappell (2000),[21] with a target DLT rate of 0.2 and a sample size of 99. We selected a larger sample size of 99 because, with smaller sample sizes, the benefits of randomization are less pronounced due to higher variability. In contrast, with a larger sample size, such as 99, randomization becomes significantly more effective. Table 5 presents the overall assessment results of accuracy and safety based on five performance metrics.

**Table 5:** Comparison of CFO and rCFO designs across different scenarios.

| Scenario | Design | MTD selection | MTD allocation | Overdose selection | Overdose allocation | Average DLT rate |
| --- | --- | --- | --- | --- | --- | --- |
| Scenario 1 | CFO | 0.6764 | 0.5083 | 0.1194 | 0.2110 | 0.1959 |
|  | rCFO | 0.6796 | 0.5120 | 0.1158 | 0.2104 | 0.1962 |
| Scenario 2 | CFO | 0.8312 | 0 | 0 | 0 | 0.3147 |
|  | rCFO | 0.8326 | 0 | 0 | 0 | 0.3149 |
| Scenario 3 | CFO | 0.7284 | 0.4816 | 0.0614 | 0.1352 | 0.1721 |
|  | rCFO | 0.7352 | 0.4806 | 0.0568 | 0.1348 | 0.1720 |
| Scenario 4 | CFO | 0.7752 | 0.5394 | 0.0154 | 0.1015 | 0.1773 |
|  | rCFO | 0.7820 | 0.5486 | 0.0160 | 0.0996 | 0.1770 |
| Scenario 5 | CFO | 0.6682 | 0.5586 | 0 | 0 | 0.1591 |
|  | rCFO | 0.6760 | 0.5640 | 0 | 0 | 0.1596 |
| Average | CFO | 0.7358 | 0.4176 | 0.0392 | 0.0895 | 0.2038 |
|  | rCFO | 0.7411 | 0.4210 | 0.0377 | 0.0890 | 0.2039 |

Simulation results indicate that by incorporating randomization, the rCFO design, with a higher value of MTD selection and allocation, marginally outperforms CFO in terms of efficiency and accuracy. In terms of safety, the rCFO design is more conservative than CFO in overdose selection and allocation, while the differences in the average DLT rate are negligible. Even small improvements in the accuracy of MTD selection and allocation can significantly impact cancer trials and new drug development, thus leading to life savings worldwide.

### 2.6 MTD selection

When completing dose assignment in a real trial, an isotonic regression is conducted on the observed DLT rates to derive the final estimates using the pool-adjacent-violators algorithm.[20] The MTD is selected as the dose level for which the isotonic estimate of the DLT rate is closest to the target rate *ϕ*. In particular, `CFO.selectmtd()` is employed for the selection of the MTD for single-drug trials, while `CFO2d.selectmtd()` is used for drug-combination trials. These two functions are adapted from **BOIN** package,[13] with some modifications. The **BOIN** package assumes a uniform prior Unif(0, 1) *≡* Beta(1, 1) for the DLT probability *p_k_*. Consequently, under the beta-binomial model, the posterior distribution of *p_k_* is Beta(*x_k_* + 1, *m_k_ − x_k_* + 1), where *x_k_* and *m_k_* respectively represent the count of DLTs and the total number of patients at dose level *k*. In contrast, for the CFO-type design, the prior for *p_k_* follows a beta distribution, Beta(*ϕ,* 1 *− ϕ*), and the corresponding posterior distribution is Beta(*x_k_* + *ϕ, m_k_ − x_k_* + 1 *− ϕ*).

As an illustration, `CFO.selectmtd()` and `CFO2d.selectmtd()` are invoked as follows:

~~~
R> ## Real trial: MTD selection for the single-drug trial
R> ntox = c(0, 0, 4, 2, 0, 0, 0); npts = c(3, 3, 27, 3, 0, 0, 0)
R> sel.single <- CFO.selectmtd(target = 0.2, ntox = ntox, npts = npts,
+ cutoff.eli = 0.95, early.stop = 0.95, verbose = TRUE)
R> summary(sel.single)
~~~

~~~
R> ## Real trial: MTD selection for the drug-combination trial
R> ntox <- matrix(c(0, 0, 2, 0, 0, 0, 2, 7, 0, 0, 0, 2, 0, 0, 0),
+ nrow = 3, ncol = 5, byrow = TRUE)
R> npts <- matrix(c(3, 0, 12, 0, 0, 3, 12, 24, 0, 0, 3, 3, 0, 0, 0),
+ nrow = 3, ncol = 5, byrow = TRUE)
R> sel.comb <- CFO2d.selectmtd(target = 0.3, npts = npts, ntox = ntox,
+ cutoff.eli = 0.95, early.stop = 0.95, verbose = TRUE)
R> summary(sel.comb)
~~~

### 2.7 Early stopping and dose elimination rules

For safety, early stopping and dose elimination rules are adopted by default for all the designs. A dose level *k* is deemed overly toxic if Pr (*p_k_ > ϕ | x_k_, m_k_ ≥* 3) *>* 0.95, where *p_k_* represents the toxicity probability of dose level *k*, and *x_k_* and *m_k_* indicate the number of observed DLTs and the number of enrolled patients at dose level *k*, respectively. If the lowest dose level is overly toxic, indicated by Pr (*p*_1_ *> ϕ | x*_1_, *m*_1_ *≥* 3) *>* 0.95, the trial will be terminated according to the early stopping rule.

Dose elimination rules are also applied to avoid assigning too many cohorts to overly toxic doses during the trial. Any dose level *k* identified as overly toxic, as well as all the higher dose levels, are eliminated. Such excluded doses are then omitted from subsequent dose allocations to prioritize patient safety. In the **CFO** package, the argument `cutoff.eli` is employed to set the threshold value for dose elimination, while `early.stop` is utilized to determine the threshold for early stopping. These thresholds can take values between 0 and 1, with higher numerical values indicating a more stringent stopping or elimination rule. Typically, the default threshold value for both early stopping and dose elimination rule is set at 0.95. This can be adjusted in practice to allow for more aggressive or conservative dose escalation strategies.

## 3 Simulations of CFO-type designs using R

The previous section outlined various CFO-type designs and how they are implemented in real trials. A single trial example may select the MTD that does not align with the target DLT due to the randomness inherent in one realized sample. Augmenting the number of enrolled patients would enrich information, leading to more robust results. However, recruiting additional patients introduces challenges such as prolonged trial duration, stringent patient eligibility criteria, and competitive recruitment. Simulations can serve as a way to evaluate how a trial design performs on average. Users can simulate the enrollment of a substantial number of patients in a single trial, prolonging the chain of dose movement to stabilize toward the target DLT. Additionally, simulations involve the synthesis of results from multiple trials conducted in parallel, assisting in the final selection. Details regarding simulations using the **CFO** package are provided in this section.

### 3.1 Evaluation metrics

The following three categories of performance metrics are used to assess the operational characteristics:

- Accuracy evaluation: This category evaluates the precision of MTD determination and allocation, encompassing the percentage of correct MTD selections (MTD selection) and the percentage of patients allocated to the MTD (MTD allocation). Higher values signify more precise and effective performance.
- Safety evaluation: This category concentrates on safety considerations, including the percentage of doses selected above the MTD (overdose selection), the percentage of patients allocated doses above the MTD (overdose allocation), and the percentage of patients experiencing DLT (average DLT rate). Smaller values indicate better safety and ethical considerations.
- Average trial duration: This metric quantifies the average duration of trials with late-onset toxicities. A shorter duration is preferable as it denotes more efficient trial execution.

In addition to the metrics above, the **CFO** package also utilizes three additional measurements to capture the distribution outcomes for multiple simulations across various dose levels, encompassing the selection percentage, the average number of observed toxicities, and the average number of treated patients at each dose level. These distributions can be visualized using the `plot()` function.

### 3.2 Execution of one simulation

In the context of simulated trials, the **CFO** package executes a single simulation of a CFO-type design using `CFO.simu()` for the CFO and aCFO design, `CFO2d.simu()` for the 2dCFO design, and `lateonset.simu()` for designs with late-onset toxicities. The argument design is used to select different designs. The function `lateonset.simu()` shares a common set of arguments with `CFO.simu()`, but with additional parameters related to late-onset outcomes (`assess.window, accrual.rate, tte.para`, and `accrual.dist`). We assign values to the time-related arguments: The DLT assessment window spans three months (`assess.window = 3`). Patients are enrolled at a rate of two per month (`accrual.rate = 2`), with their arrival times following a uniform distribution (`accrual.dist = ‘unif’`). The occurrence of DLT events is simulated using a Weibull distribution. The two parameters of the Weibull distribution are determined by the proportion of DLTs occurring during the first half of the assessment window (`tte.para`) and the proportion of DLTs occurring within the entire assessment window, which is fixed as `p.true`. Users have the flexibility to adjust the argument tte.para according to specific needs, with tte.para set to 0.5 in this example.

Taking the CFO, f-aCFO and 2dCFO designs as examples, the following code executes the design, displaying the output in a textual summary and plotting the trajectory of dose level movements (Figure 3).

**Figure 3:**
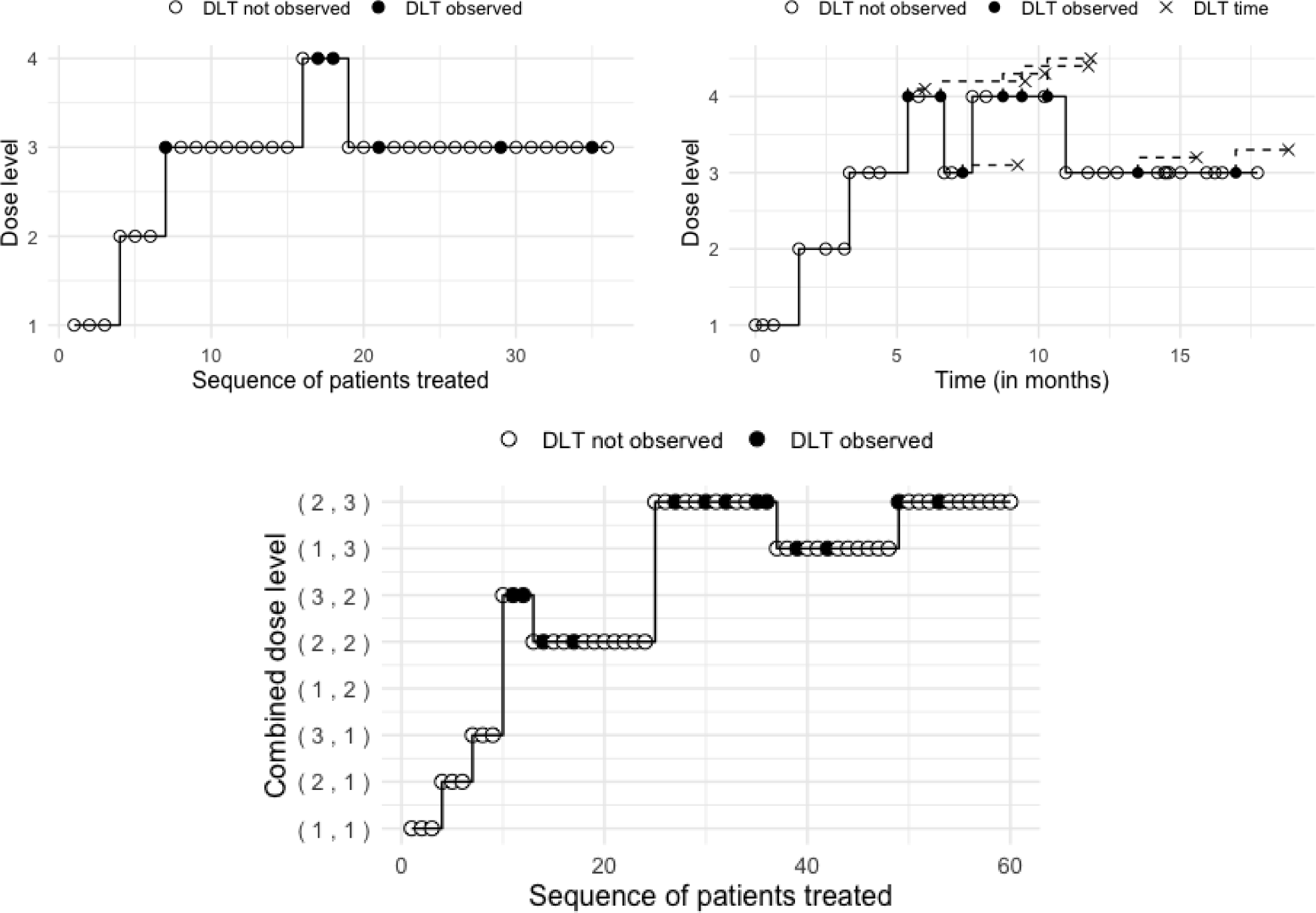
Illustration of a hypothetical trial using the CFO (top left), f-aCFO (top right), and 2dCFO (bottom) designs. Patients are treated in a cohort size of 3, where solid circle *•* and empty circle *◦* respectively indicate the presence or absence of observed toxicity in patients, and the x-axis value of the cross *×* signifies the time at which the DLT occurred.

~~~
R> ## Execution of a single CFO simulation
R> p.true <- c(0.01, 0.07, 0.20, 0.35, 0.50, 0.65, 0.80)
R> target <- 0.2; prior.para <- list(alp.prior = target, bet.prior = 1 - target)
R> CFOtrial <- CFO.simu(design = ‘CFO’, target = 0.2, p.true = p.true,
+ init.level = 1, ncohort = 12, cohortsize = 3, prior.para = prior.para,
+ cutoff.eli = 0.95, early.stop = 0.95, seed = 1)
R> summary(CFOtrial)
R> plot(CFOtrial)
~~~

~~~
R> ## Execution of a single f-aCFO simulation
R> faCFOtrial <- lateonset.simu (design = ‘f-aCFO’, target = 0.2, p.true = p.true,
+ init.level = 1, ncohort = 12, cohortsize = 3, assess.window = 3,
+ tte.para = 0.5, accrual.rate = 2, accrual.dist = ‘unif’,
+ prior.para = prior.para, cutoff.eli = 0.95, early.stop = 0.95, seed = 1)
R> summary(faCFOtrial)
R> plot(faCFOtrial)
~~~

~~~
R> ## Execution of a single 2dCFO simulation
R> p.true <- matrix(c(0.05, 0.10, 0.15, 0.30, 0.45, 0.10, 0.15, 0.30, 0.45, 0.55,
+ 0.15, 0.30, 0.45, 0.50, 0.60), nrow = 3, ncol = 5, byrow = TRUE)
R> CFO2dtrial <- CFO2d.simu(target = 0.3, p.true = p.true, init.level = c(1,1),
+ ncohort = 20, cohortsize = 3, cutoff.eli = 0.95, early.stop = 0.95, seed = 1)
R> summary(CFO2dtrial)
R> plot(CFO2dtrial)
~~~

Compared to a single-drug trial, the drug-combination trial involves a larger number of dose-level combinations. Achieving the target dose level requires more moves, necessitating the enrollment of a larger number of patients to ensure accuracy. Twelve cohorts are enrolled for single-drug trials while 20 cohorts are enrolled for the drug-combination trial. After a brief fluctuation, the dose level ultimately converges to dose level 3 for the CFO design, dose level 3 for the f-aCFO design, and the dose level combination (2, 3) for the 2dCFO design. These outcomes are all aligned with the target DLTs.

### 3.3 Operating characteristics evaluation with multiple simulations

Extensive simulations are often needed for evaluating the operational characteristics of trial designs. With `CFO.simu()`, `lateonset.simu()` and `CFO2d.simu()` as intrinsic components, the `CFO.oc()` and `CFO2d.oc()` functions are employed to conduct extensive simulations and obtain the operational characteristics for single-drug and drug-combination trials, respectively. The argument nsimu represents the number of simulations, and seeds is a vector containing random seeds for each simulation. As the CFO and aCFO designs do not involve late-onset outcomes, the time-related arguments (`assess.window, accrual.rate, tte.para`, and `accrual.dist`) are set to `NA`. For designs with late-onset toxicity, specific values are assigned to time-related arguments, replacing `NA`. The code below specifies these configurations and then conducts multiple simulations for the CFO, f-aCFO, and 2dCFO, respectively.

~~~
R> ##Multiple simulations for the CFO design
R> p.true <- c(0.01, 0.07, 0.20, 0.35, 0.50, 0.65, 0.80)
R> target <- 0.2; prior.para <- list(alp.prior = target, bet.prior = 1 - target)
R> CFOoc <- CFO.oc(nsimu = 5000, design = ‘CFO’, target = 0.2, p.true = p.true,
+ init.level = 1, ncohort = 12, cohortsize = 3, assess.window = NA,
+ tte.para = NA, accrual.rate = NA, accrual.dist = NA, prior.para = prior.para,
+ cutoff.eli = 0.95, early.stop = 0.95, seeds = 1:5000)
R> summary(CFOoc)
~~~

~~~
R> ##Multiple simulations for the f-aCFO design
R> faCFOoc <- CFO.oc(nsimu = 5000, design = ‘f-aCFO’, target = 0.2,
+ p.true = p.true, init.level = 1, ncohort = 12, cohortsize = 3,
+ assess.window = 3, tte.para = 0.5, accrual.rate = 2,
+ accrual.dist = ‘unif’, prior.para = prior.para, cutoff.eli = 0.95,
+ early.stop = 0.95, seeds = 1:5000)
R> summary(faCFOoc)
~~~

~~~
R> ##Multiple simulations for the 2dCFO design
R> p.true <- matrix(c(0.05, 0.10, 0.15, 0.30, 0.45, 0.10, 0.15, 0.30, 0.45, 0.55,
R> 0.15, 0.30, 0.45, 0.50, 0.60), nrow = 3, ncol = 5, byrow = TRUE)
R> target <- 0.3; prior.para <- list(alp.prior = target, bet.prior = 1 - target)
R> CFO2doc <- CFO2d.oc(nsimu = 1000, target = 0.3, p.true = p.true,
+ init.level = c(1,1), ncohort = 20, cohortsize = 3, prior.para = prior.para,
+ cutoff.eli = 0.95, early.stop = 0.95, seeds = 1:1000)
R> summary(CFO2doc)
~~~

The `summary()` function offers information on the count of early stopping occurrences in simulations along with the values of performance metrics. Below is a textual description provided by summary(faCFOoc).

~~~
In 5000 simulations, early stopping occurred 13 times.
Among simulations where early stopping did not occur:
Selection percentage at each dose level:
0.000 0.146 0.606 0.224 0.019 0.001 0.000
Average number of patients treated at each dose level:
3.404 7.478 12.863 8.171 3.056 0.825 0.122
Average number of toxicities observed at each dose level:
0.037 0.530 2.582 2.865 1.510 0.537 0.097
Percentage of correct selection of the MTD: 0.606
Percentage of patients allocated to the MTD: 0.358
Percentage of selecting a dose above the MTD: 0.244
Percentage of allocating patients at dose levels above the MTD: 0.339
Percentage of the patients suffering DLT: 0.227
Average trial duration: 20.5
~~~

In addition, the function `plot()` provides the distribution for better visualizing the output. Figures in Appendix B illustrate the MTD selection rate, average patient allocation, and average DLT observed at different dose levels, depicting the CFO, f-aCFO, and 2dCFO designs’ performance.

## 4 Discussion

Due to the nonparametric or model-free nature as well as utilization of information on multiple doses for decision making, CFO-type designs are regarded as robust, efficient, and easy-to-use approaches for conducting phase I trials. Another key distinction of CFO from existing methods is that it only requires the specification of the target toxicity rate, a minimal component in the design specification, hence referred to as “calibration-free”. The **CFO** package, as a user-friendly and well-documented tool, provides a comprehensive implementation of various CFO-type designs. It covers all key aspects of the phase I clinical trial design, including the determination of dose levels for substantial cohorts, selection of MTD for a real trial, and execution of single or multiple simulations to obtain operating characteristics. The distinctive feature of **CFO** is its flexibility in accommodating diverse CFO-type designs, allowing users to tailor the approach based on factors such as dose information inclusion, handling of late-onset toxicity, and the nature of the target drug (single-drug or drug-combination). This adaptability enhances the applicability of **CFO** to a wide range of clinical trial scenarios.

Beyond its functional depth, **CFO** stands out by providing user-friendly assessment tools, offering both descriptive and graphical outputs. This user-centric approach is valuable, providing researchers with an intuitive understanding of the design’s operational dynamics and outcomes. Consequently, clinical trial statisticians can easily present results to clinicians and facilitate discussion about the appropriateness of the design. This tutorial aids users in grasping the package’s functionalities, enhancing its effective utilization.

## Data Availability

All data produced in the present work are contained in the manuscript

## Appendices

### Appendix A Package Details

#### A.1 Overview of the user-visible functions in the CFO package

Table 6 is an overview of available functions in the **CFO** package. Users can refer to the documentation (e.g., `help(“CFO.next”)`) for function arguments and detailed return types.

**Table 6:**
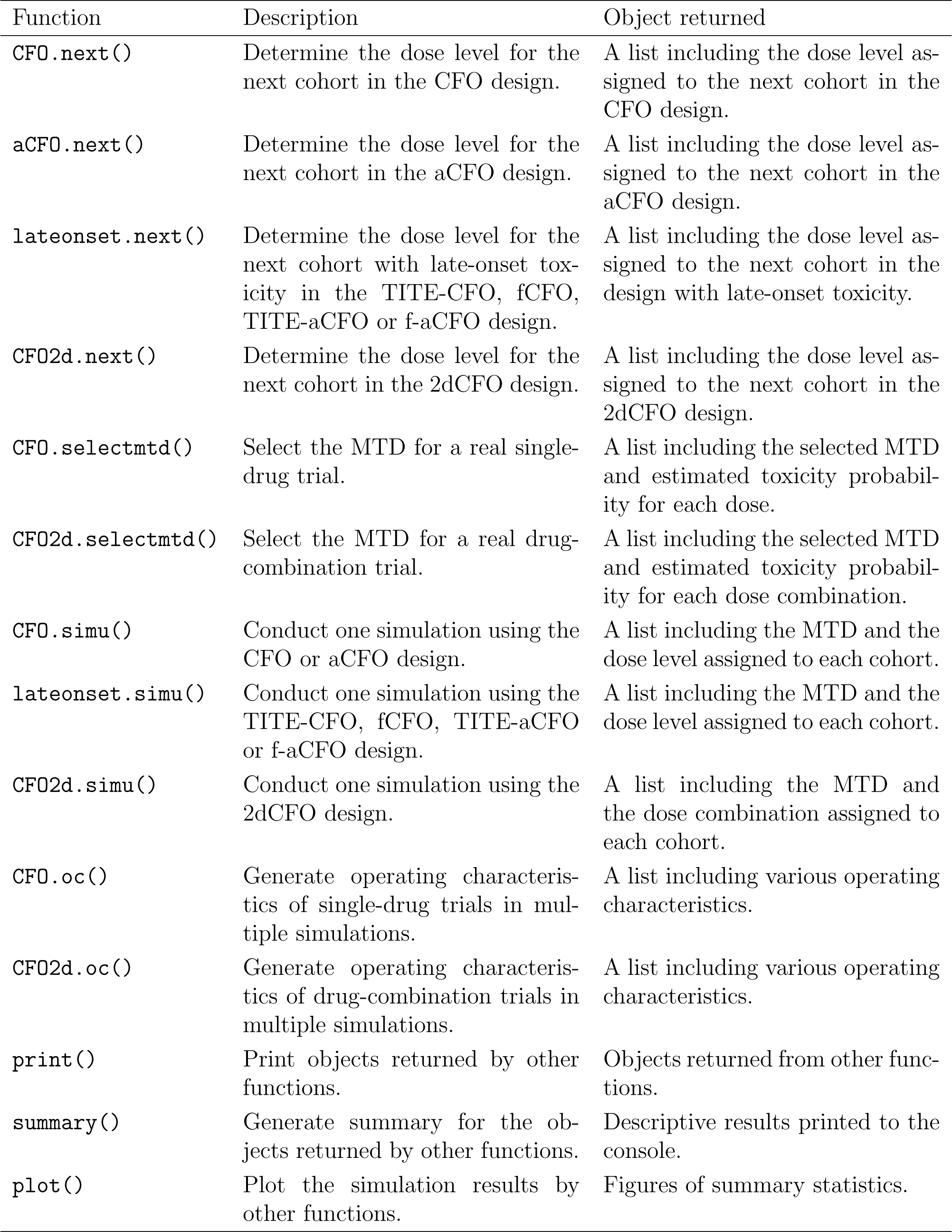
Overview of the user-visible functions in the CFO package.

#### A.2 Descriptions of the arguments in the CFO package

Table 7 shows the details of the arguments used in the functions of the **CFO** package.

**Table 7:**
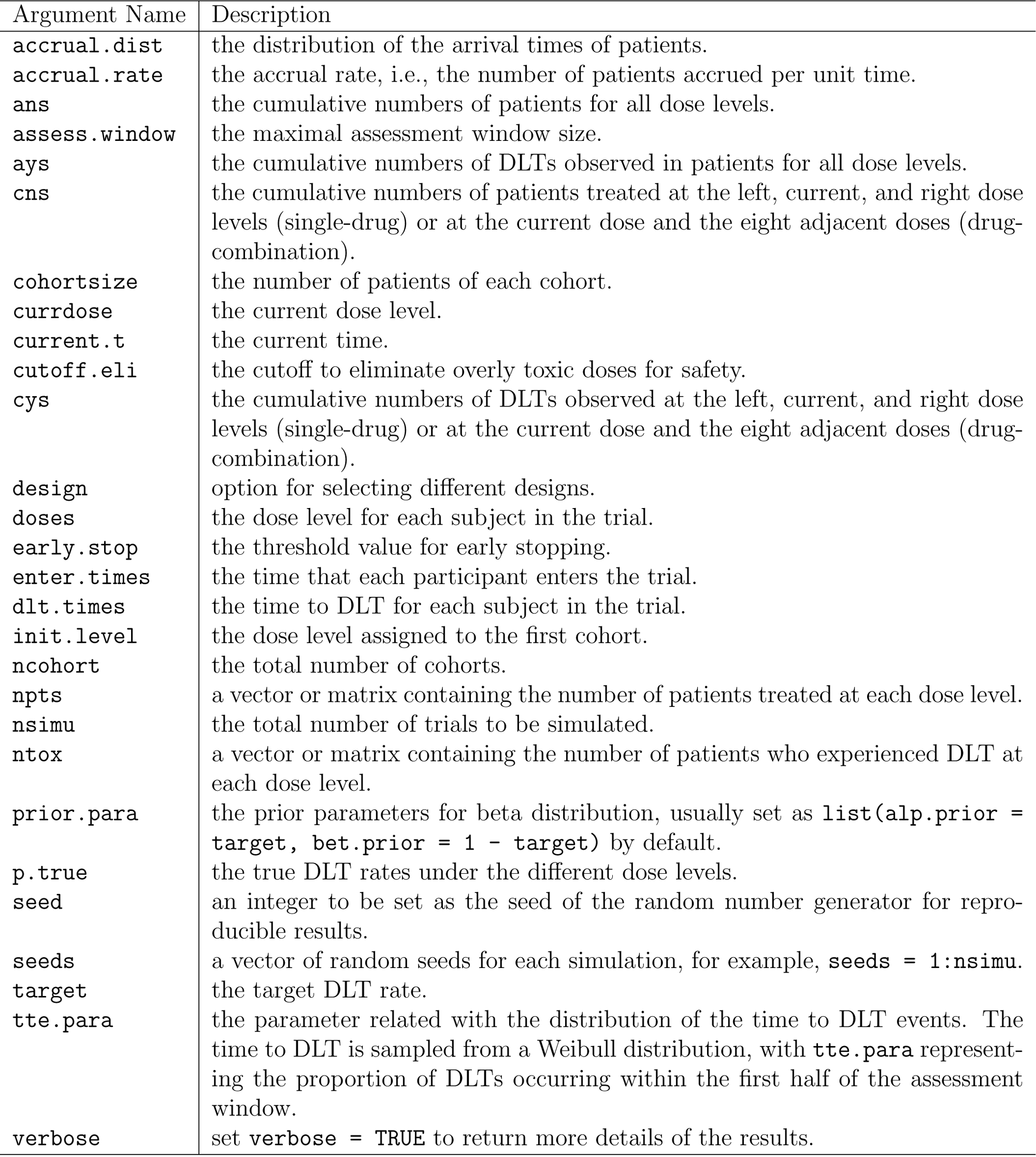
Descriptions of the arguments in the CFO package.

### Appendix B Multiple simulation results

**Figure 4:**
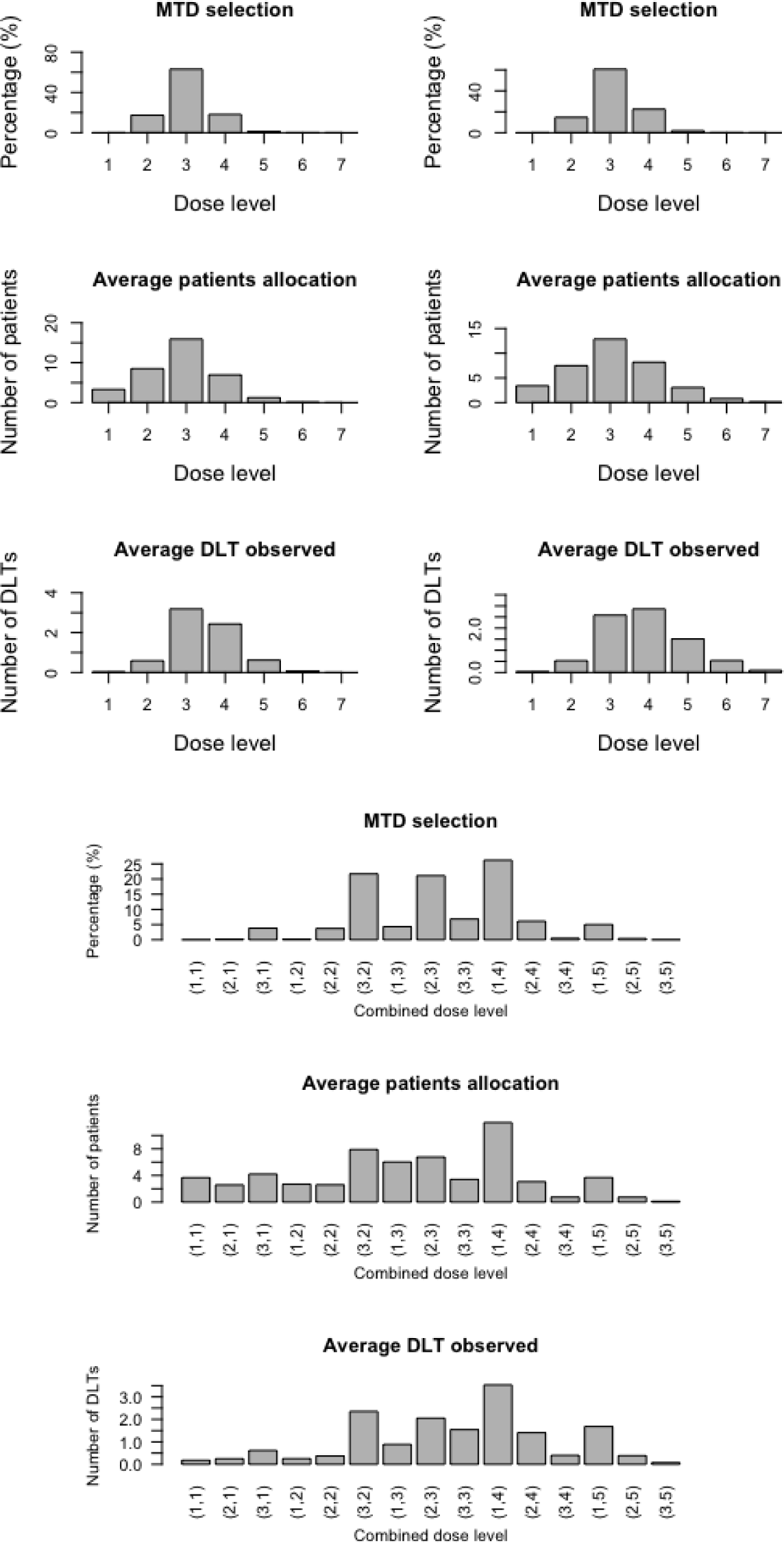
Operating characteristics obtained from 1000 simulations using the CFO (top left), f-aCFO (top right) and 2dCFO (bottom) design.

